# Deep Sequence Learning for Assessing Hypertension in Pregnancy from Doppler Signals

**DOI:** 10.1101/2022.01.26.22269921

**Authors:** Nasim Katebi, Gari D. Clifford

## Abstract

Measuring blood pressure during pregnancy is an essential component of antenatal care, and is critical for detecting adverse conditions such as pre-eclampsia. The standard approach for measuring blood pressure is via manual auscultation by a trained expert or via an oscillometric self-inflating cuff. While both methods can provide reasonably accurate blood pressure measurements when used correctly, non-expert use can lead to significant error. Moreover, such techniques are uncomfortable and can cause bruising, pain and consequential resistance to use / low compliance. In this work, we propose a low-cost onedimensional Doppler-based method for the detection of hypertension in pregnancy.

Using a sample of 653 pregnant women of Mayan descent in highland Guatemala, we recorded up to 10 minutes of 1D Doppler data of the fetus, and blood pressure from both arms using an Omron M7 oscillometric cuff. A hierarchical LSTM network with attention mechanism was trained to classify hypertension in pregnancy, producing an area under the receiver-operator curve of 0.94. A projection of the data into lower dimensions indicates hypertensive cases are located at the periphery of the distribution of the output of the distribution.

This work presents the first demonstration that blood pressure can be measured using Doppler (without occlusion) and may lead to a novel class of blood pressure monitors which allow rapid blood pressure estimation from multiple body locations. Moreover, the association of the predictor with the fetal blood flow indicates that hypertension in the mother has a significant effect on the fetal blood flow.

## Introduction

Hypertension is the most common medical complication encountered during pregnancy,y where 10% of women experience blood pressure above normal during pregnancy. Preeclampsia is characterized by high blood pressure and is a major cause of maternal and perinatal morbidity mortality that complicates 2% to 8% of pregnancies (1, 2). Preeclampsia is a pathological condition in pregnancy initiated by abnormal uteroplacental hemodynamics (3). Placentation abnormalities lead to the symptomatic stage, wherein the pregnant woman develops hypertension, defined as at least two repeated blood pressure measurements greater than or equal to 140 mmHg systolic blood pressure (SBP) or 90 mmHg diastolic blood pressure (DBP) (4).

One important aspect of diagnosing and managing hypertension in pregnancy is early identification of pregnancies at high-risk of early-onset pre-eclampsia and undertaking the necessary measures to improve placentation and reduce the prevalence of the disease (5). Adverse outcomes related to hypertensive disorders of pregnancy can affect both mother and fetus in long- and short-term. The adverse effects are associated with placental abruption, preterm delivery, fetal growth restriction, stillbirth, maternal death secondary to stroke and eclampsia, as well as future risk of hypertension, diabetes mellitus, and cardiovascular disease in the mother (6, 7). This highlights the importance of accurate monitoring of blood pressure during antenatal care and motivates us to design the study on the detection of hypertension in pregnancy.

Both Fetal heart rate variability (FHRV) and fetal ECG morphology have been shown to be diagnostic biomarkers for mild and severe pre-eclampsia. Yum et al. (8) conducted the study on the instability and frequency-domain variability of fetal heart rate in three study groups of control, severe pre-eclampsia and severe pre-eclampsia affected by intrauterine growth restriction (IUGR) all underwent routine followups at Samsung Medical Center. Results demonstrated that low- and high-frequency power were significantly higher in the group not affected by IUGR when compared to the control group. However, in the group affected by IUGR, low-frequency power was significantly lower and high-frequency power was not significantly different in comparison to the control group. Another study was presented by Lakhno on the effect of pre-eclampsia on FHRV (9). In this study, the modulated fetal CTG variables captured the suppression of fetal biophysical activity and the development of fetal distress in severe pre-eclampsia. In addition to the impact of pre-eclampsia on FHRV indices, Lakhno studied changes in fetal ECG morphology. The presented results revealed that FHRV metrics were directly related to the severity degree of pre-eclampsia. Also in pre-eclampsia cases shortening of PQ and QT and increased T/QRS ratio were observed (10). In addition, alterations in resistance and flow could lead to a chamber remodeling during early development. Aye et al. (11) performed a study on prenatal and postnatal cardiac development in fetuses born to either pre-eclampsia or gestational hypertension. This study demonstrated that term-born infants from hypertensive pregnancies had persistently smaller right ventricular end-diastolic volumes. At 3 months of postnatal life, infants born to hypertensive pregnancies also showed subtle changes in left ventricular mass. These findings were similar to those presented by Timpka et al. (12) which showed that fetuses born to pre-eclamptic pregnancies and gestational hypertension had greater left ventricular relative wall thickness, with smaller left ventricular end-diastolic volumes.

One non-invasive method for capturing fetal cardiac activity is a one-dimensional Doppler ultrasound (1D-DUS) with the advantage of providing a low-cost and simple method for fetal heart rate monitoring. The Doppler transducer transmits and receives ultrasound waves, which reflect the fetal cardiac activity. Using the 1D-DUS signal, blood flow, cardiac wall and valve motions can be captured, and they are differentiable based on their different velocities. Figure 1 shows the simultaneous ECG and 1D-DUS signal. A review of using 1D-DUS to assess vascular changes in pre-eclampsia indicates the effectiveness of deriving discriminating parameters from this data to diagnose pregnancy complications (13). Also, there are multiple parameters derivable from the 1D-DUS signals which are related to placental perfusion, including resistance index, pulsatility index, or systolic/diastolic ratio from uterine artery (14, 15), fetal heart rate responses and uteroplacental flows (16). Due to the advantages of 1D-DUS in both the recording technique and assessment of fetal wellbeing, we designed a predictive model using fetal 1D-DUS recordings to detect hypertension.

**Fig 1.**
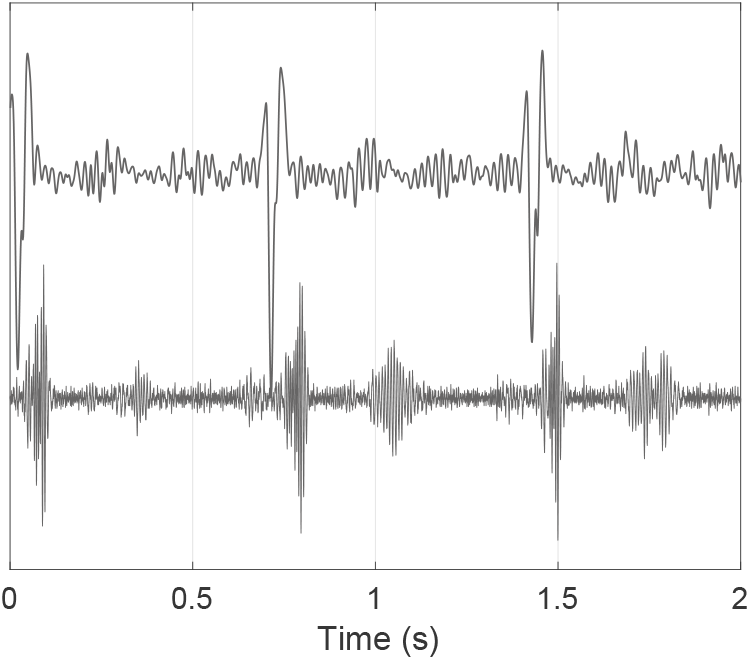
Fetal Doppler ultrasound and simultaneously recorded fetal electrocardiogram: (top) fetal ECG signal and (bottom) fetal Doppler signal.

Recent advances in deep learning, especially recurrent neural network (RNN) (17) and long short-term memory (LSTM) (18) models provide useful insights on how to tackle the problems relating to sequence modeling, time series classification and prediction. LSTM have been broadly applied to time series data analysis due to its capability in processing of long sequences of data. Given that we have a sequential data, it is natural to consider the use of a recurrent neural network to keep track of the variability and temporal dependency. Attention mechanisms have been shown to be effective in improving the performance of sequence learning models by attending to every hidden state and then making predictions after deciding which one is more informative (19, 20). Various studies showed the effect of using appropriate loss function in model performance. The contrastive loss has been shown to perform well in self-supervised algorithms. The supervised contrastive learning approach presented by Kholsa et al. (21) is based on pulling the normalized embeddings from the same class closer together than embeddings from different classes, which leads to achieving discriminative features with smaller inter-class variability (21, 22). Our intent in this study is to discover the relation between fetal cardiac activity and maternal blood pressure to detect hypertensive cases. The approach taken to the blood pressure estimation used in this work is based upon a hierarchical attention network (23), which is a stable and powerful method for extracting features from sequential data and learn short- and long-range dynamics. This approach is used to model time dependencies in fetal 1D-DUS signal and capture the variability of the cardiac activity. In addition, multiple steps were taken to deal with the non-uniform distribution of blood pressure levels and in particular, the under-representation of hypertensive subjects. The following sections detail the approach to dealing with these problems, with a focus on identifying hypertension in pregnancy.

## Methods

### Data Sources

In this work, we used 1D-DUS recordings from 653 pregnant women (736 visits) at 5 to 9 months of gestation plus contemporaneous blood pressure measurements from the Guatemala RCT Database, which were collected as part of a randomized control trial, conducted in rural highland Guatemala in the vicinity of Tecpan, Chimaltenango (24, 25). The study focused on the impact of a mHealth decision support system to improve the continuum of care for indigenous women of the target region. It was approved by the Institutional Review Boards of Emory University, the Wuqu’ Kawoq | Maya Health Alliance, and Agnes Scott College (Ref: IRB00076231 - ‘Mobile Health Intervention to Improve Perinatal Continuum of Care in Guatemala’) and registered as a clinical trial (ClinicalTrials.gov identifier NCT02348840). This data set includes 1D-DUS signals, recorded by traditional birth attendants (TBAs), who were trained to use the hand-held 1D-DUS device. Immediately before recording the 1D-DUS signals, the TBA also entered the gestational age in months and the maternal heart rate, and captured maternal blood pressure by photographing the screen of a self-inflating blood pressure device. The blood pressure monitor device used was an Omron M7 (Omron Co., Kyoto, Japan), which has been validated in a preeclampsic population (26). The blood pressure values were extracted from the images, and the transcription method was designed to automatically detect the LCD in the photo and extract the SBP and DBP values (27). The 1D-DUS device was an AngelSounds Fetal 1D-DUS JPD-100s (Jumper Medical Co., Ltd., Shenzhen, China) with an ultrasound transmission frequency of 3.3 MHz. Data were captured at 44.1 kHz, using a Samsung S2, S3 mini or S4 mini and stored as uncompressed WAV files at 7056 Kb/s (16 bits). All data were captured using the same mobile application designed to record the 1D-DUS. Figure 3-a illustrates the devices used in this study.

Blood pressure was recorded in a supine position which is slightly lower than blood pressure in a sitting position. The BP difference by body position is estimated as mean SBP difference=4.02 mmHg and DBP=2.97 mmHg (28). Figure 2 shows the joint distribution of the average left-right arms SBP and DBP after adjustment for supine position. In this dataset, there are 3 severe hypertensive cases (SBP>160 or DBP>110 mmHg) and 7 mild/moderate hypertensive cases (SBP 140-160 or DBP 90-110 mmHg) recordings. And, table 1 shows the mean and standard deviation of recorded blood pressure based on gestational age.

**Table 1.** Mean and standard deviation of blood pressure (mmHg) in gestational ages 5 to 9 months in the dataset.

| Gestational age | 5 | 6 | 7 | 8 | 9 |
| --- | --- | --- | --- | --- | --- |
| $mean_{SBP}$ | 99.6 | 99.4 | 101.5 | 103.1 | 109.3 |
| $std_{SBP}$ | 8.0 | 6.9 | 8.7 | 9.0 | 13.5 |
| $mean_{DBP}$ | 60.1 | 60.1 | 62.9 | 62.4 | 69.5 |
| $std_{DBP}$ | 7.6 | 6.5 | 5.9 | 7.3 | 9.7 |

**Fig 2.**
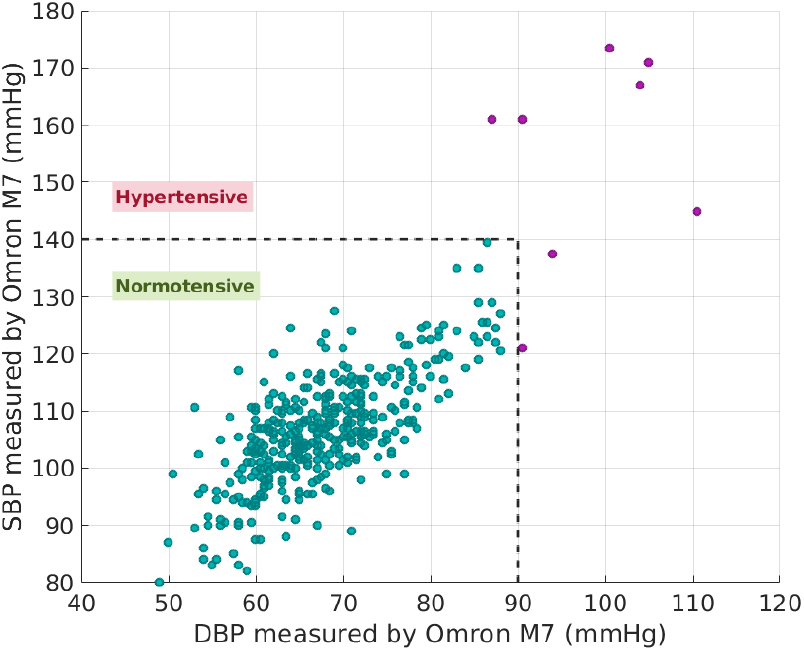
Joint distribution of average left-right arm SBP and DBP of all patients after adjustment for supine position used in this work. Observations with SBP greater than or equal to 140 mmHg or DBP 90 mmHg were considered as hypertensive cases.

### Overview of algorithms

Our method for detecting hypertension in pregnancy from fetal 1D-DUS signals is structurally similar to that used in (23). In order to improve the representation learning, we used the training approach presented by Khosla et al. (21). Given the input signal, the sequence of scalogram images were generated, then the hierarchical attention network was used to encode the sequence and obtain the normalized embedding. The representation was further propagated through a projection network that is discarded in inference time. The encoder and projection networks were trained by using a contrastive loss function. The weights of the encoder network was then freezed and used on top of the classification network to classify hypertension. Figure 3-(b,c) illustrates the overview of the model and the training process.

**Fig 3.**
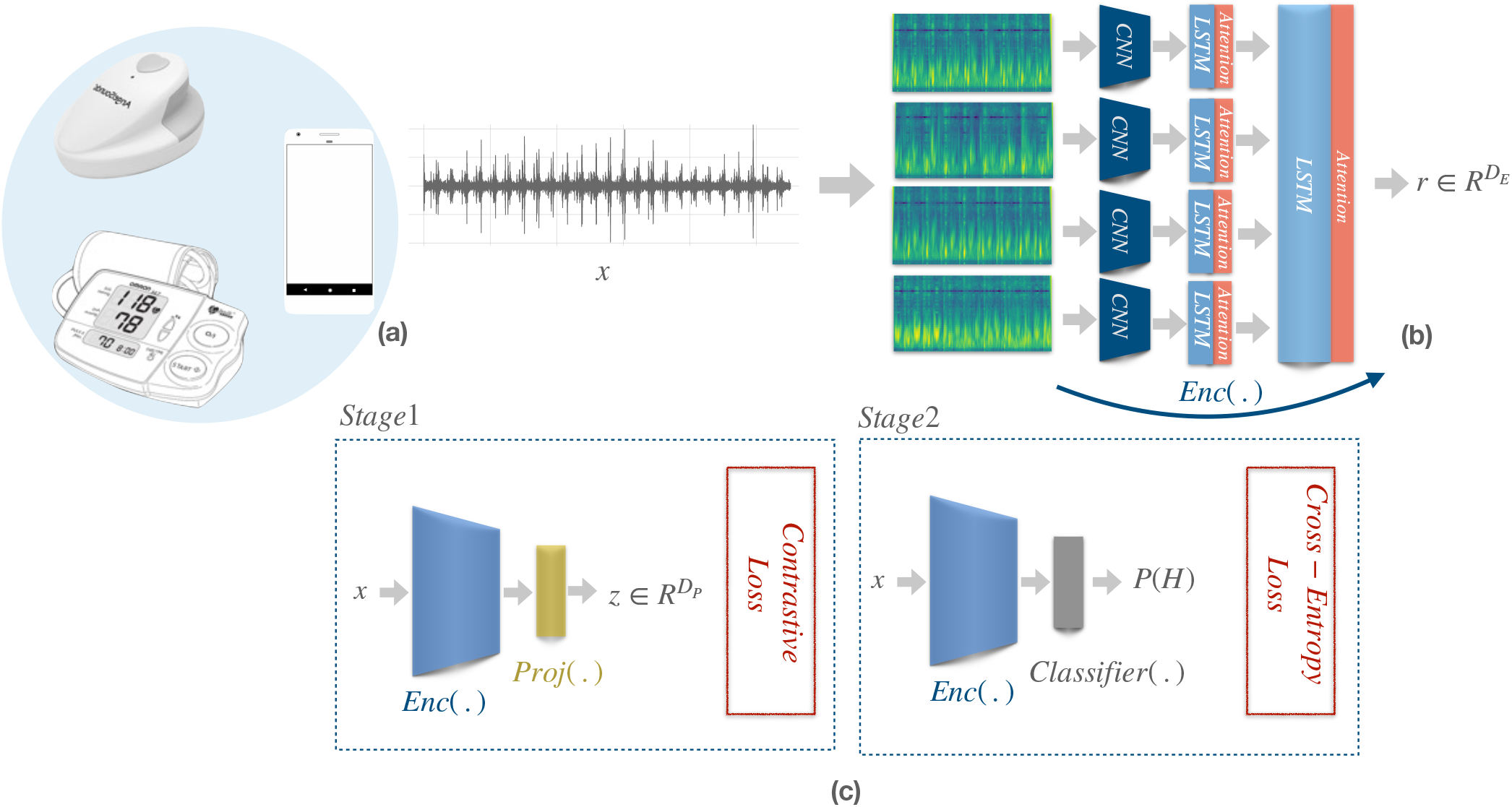
An overview of the proposed process for training a deep sequence learning model to detect high maternal blood pressure from abdominal Doppler acquired during routine fetal monitoring. a) Data sources. The 1D ultrasound is captured by a cellphone in rural communities by a TBA using Doppler transducer. The blood pressure is captured using an oscillometric cuff device and the result is recorded on the phone via. photograph taken by the TBA. The DUS and photo are uploaded to AWS and the image is transcribed into a BP reading. b) The scalogram of 1D-DUS is calculated and fed to hierarchical attention network. c) The mapping is learnt through two step training process including stage 1, representation learning by updating weights of encoder using contrastive loss and stage 2, freezing the encoder and training the classifier to estimate the probability of hypertension (*P* (*H*)).

### Supervised Contrastive learning

The idea of supervised contrastive learning is to teach the network to learn how to map the normalized encoding of samples belonging to the same category closer and the samples belonging to the other classes farther. In the deep sequence classification models, the network converts the signals into a representation and then uses these representations to classify the signals. So forming the representation using the contrastive learning approach leads to having better performance in the classification stage. The components of the approach used in this study are as follows:

- *Sequence Encoder, Enc(*.). The *Enc*(.) network consists of two levels of LSTM structure. Processing in LSTM is realized by three key gate units: input gate, output gate, and forget gate, which are used for implementing information protection and control. In order to process the 1D-DUS signal, two-step sequence modeling was leveraged. The attention network in both steps can assign larger weight to the most important sections of the input data regarding the objective of the problem. The *Enc*(.) network, maps input *x* to a representation vector 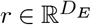. *r* is normalized to the unit hypersphere in 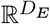.
- *Projection Network, Proj(*.). The projection network is a multi-layer perceptron and maps *r* to a vector 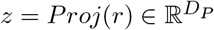. The output was again normalized to lie on the unit hypersphere, which enables using an inner product to measure distances in the projection space (stage 1 of training).
- *Classifier Network, Classifier(*.). The classifier network is also a multi-layer perceptron. In stage 2 of training process, the embeddings from the *Enc*(.) network is fed to the classifier network and cross-Entropy loss is used to optimize the parameters.

### Experimental set-up

#### 1D-DUS signal processing

Given the nature of the physiological time-series, 1D-DUS signals are likely to be corrupted with internal and external interference such as respiration, movement, and environmental noise. In this work, a second-order band-pass Butterworth filter was used to reduce the noise. By observing the frequency components of the 1D-DUS signals, the cut-off frequencies were set to 25 and 600 Hz, corresponding to cardiac oscillations. In addition, the signal quality assessment method presented in (29) was used before processing to exclude low quality recordings. After the quality assessment, each 5 minutes recording was devided into 3.75 seconds and the scalogram was generated using 50 ms Hanning window for better representation of the signal in time and frequency.

#### Network implementation

The sequence encoder network (*Enc*(.)) gets the scalogram of the signal and consists of three layers of 2-D convolutional neural network (kernel size=(3,3)). Each layer is followed by batch normalization, rectified linear (ReLU) units, and max pooling units. Then the extracted feature was fed to the first step of sequence encoder networks which consist of LSTM networks with 50 units. The projection network *proj*(.) is one dense layer with 32 units and the classification layers includes dropout layer and dense layer with 32 units. For the representation learning step, the contrastive learning loss function and for the classification part, the cross-Entropy loss function was used. Mini batch stochastic gradient descentt (SGD) was leveraged to optimaze the parameters of the network. The proposed method is implemented in tensorflow 2.0 and Python3.

#### Evaluation metrics

The network was trained and tested using five minutes 1D-DUS recordings. To evaluate the model performance, confusion matrix, Receiver operating characteristic (ROC) and area under ROC curve (AUC) were provided. The mapping of data in two dimensional space is also provided to show the performance of encoder network.

## Results

### Classification results

Figure 4 shows the performance of the classification model on randomly sampled test data. This resulted to 180 true negative, 1 false positive, 2 true positive and 2 false negative.

**Fig 4.**
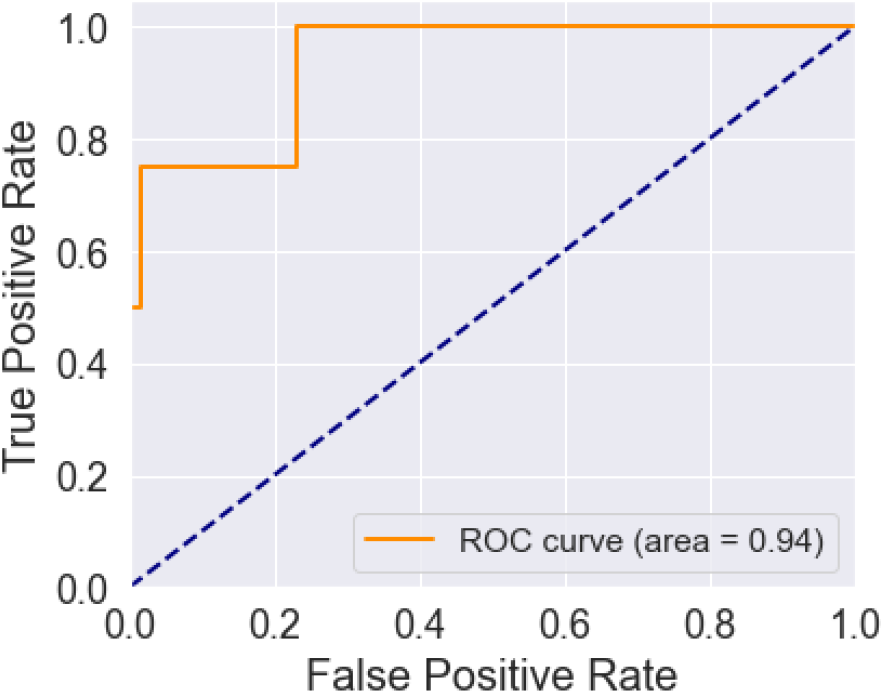
Result of classification of normotensive and hypertensive pregnancies. a) Receiver operating characteristic and b)confusion matrix.

### Regression results

We also tested the performance of the model on blood pressure estimation. Therefore, after the representation learning the output of the encoder network was used to train the regression model. Figure 5 shows the result of SBP estimation from 5 minutes 1D-DUS signals.

**Fig 5.**
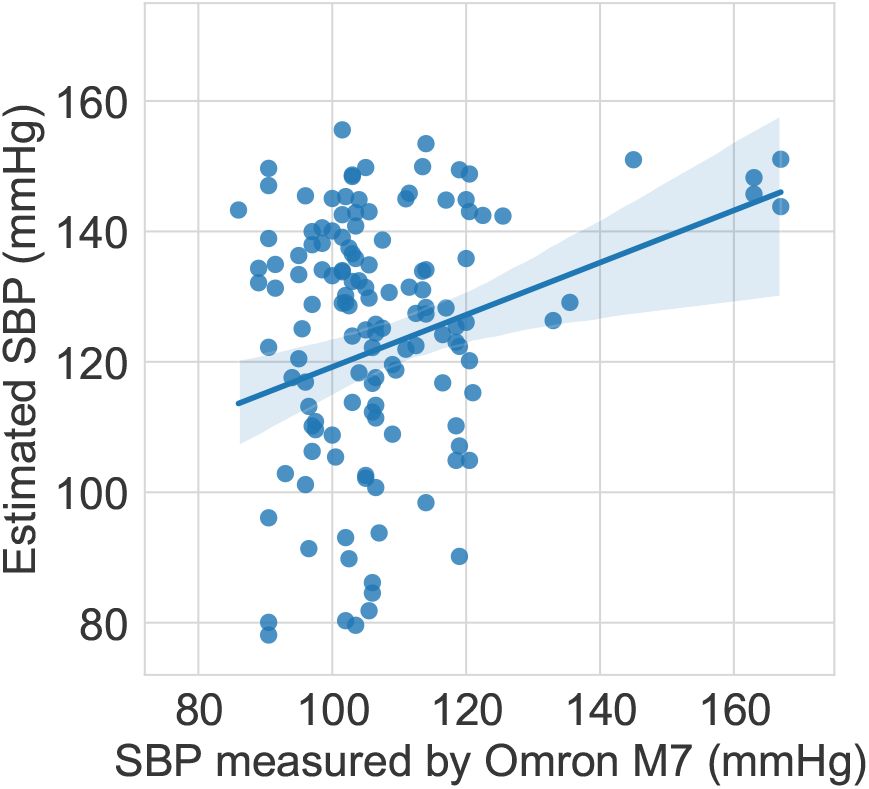
Blood pressure estimation from 5 minutes Doppler signals.

### Qualitative results

Figure 6 shows the sample test data in two-dimensional space. In order to visualize the data in lower space, t-Distributed Stochastic Neighbor Embedding (tSNE) method was applied on the output of *proj*(.) layer. The mapping presented in here indicates hypertensive cases are located at the periphery of the distribution. The data embedding is provided for the set of train and test data (figure 6).

**Fig 6.**
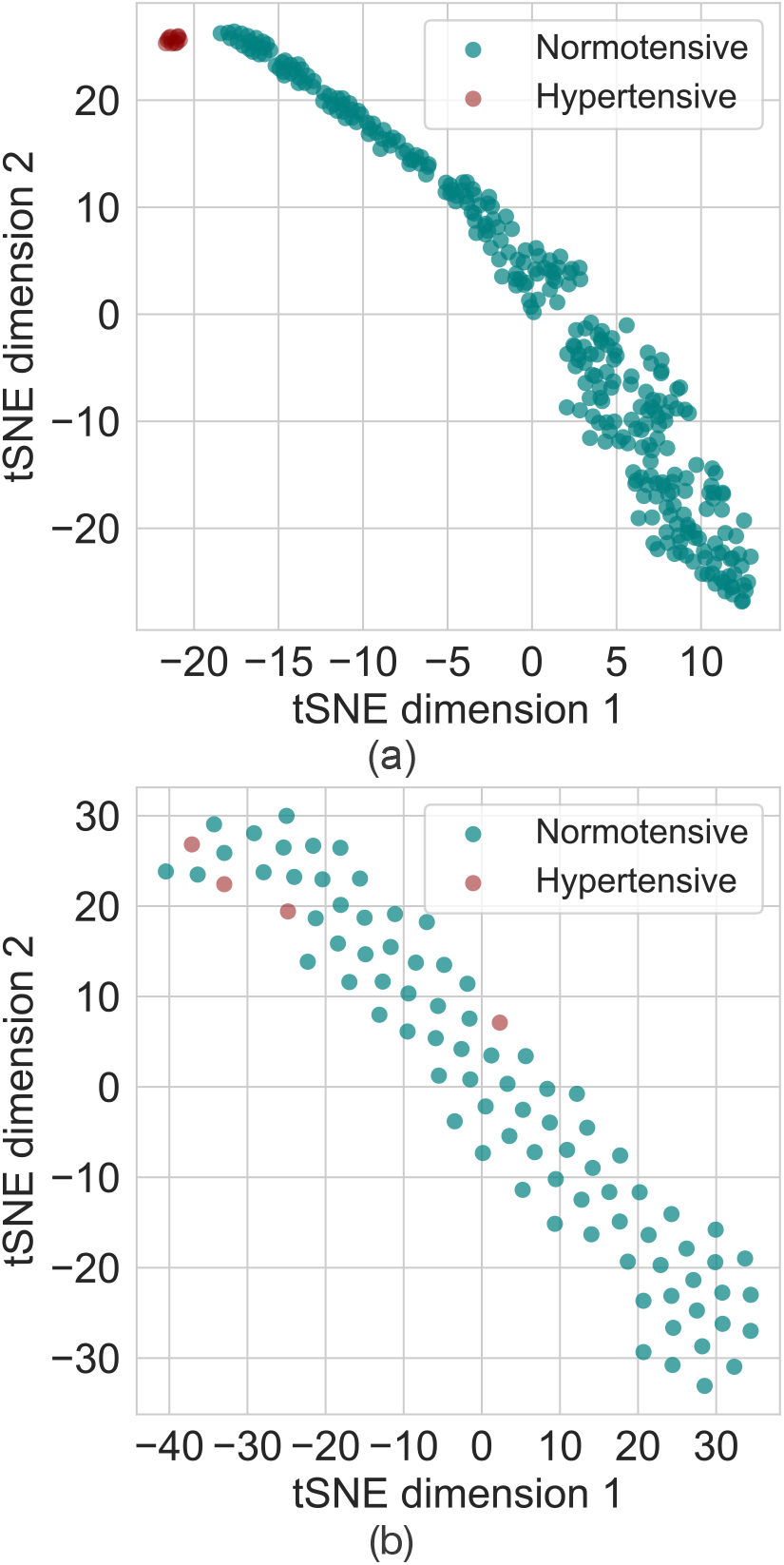
Training data (a) and Test data (b) represented in a two dimensional space. The dimension was reduced to two by applying the tSNE method on the output of the projection layer. Note that the data associated with the hypertensive patients are located at the extremes.

Note that the small number of hypertensive examples leads to a slight overfitting, although clearly the hypertensive events are still located at one extreme of the projected distribution. We can therefore infer that the model can learn the discriminative features in the training phase, and the overfiting can likely be mitigated by adding more samples of high blood pressure cases.

## Discussion

The results presented here demonstrate that the classification accuracy and representation learning respectively provide sufficiently accurate hypertension detection. We note several minor limitations of the current study. First, there were relatively few hypertensive subjects in our cohort (3 severe hypertension and 7 hypertension in totall of 736 visits) and the subjects were all single race (Native Central American of Mayan origin from rural highland Guatemala). Although this is initially a strength, and helps reduce variables associated with race and environment, it prevents a definitive claim that this would extend to other races and settings. Never-the-less, we see no principled reason that the approach would not extend beyond the population studied (and indeed, the medical community has used blood pressure monitors largely developed on mostly Caucasian cohorts for decades without significant objections from the regulatory authorities or medical community.) As we continue to collect data, we will continue to expand the cohort for a range of diverse conditions, blood pressure ranges and populations.

It is important to note that blood pressure has been estimated from Doppler before. However, when measuring blood pressure with a Doppler, the principle is to occlude arterial blood flow by inflating a cuff and then deflating it until the flow goes back to normal. When the pressure in the cuff is just below the systolic blood pressure, blood flow can pass the cuff and is detected by the Doppler probe. This ‘sphygmomanometry’ approach requires significant additional equipment and is much less pleasant for the patient. Moreover, it cannot be easily integrated into the routine monitoring process for ultrasound screening of fetuses.

The presented model for hypertension detection is based on the processing of fetal cardiac activity, therefore, it is important to consider the effect of hypertension on fetus and possible cases of IUGR. In this study, clinical labels of preeclampsia and IUGR conditions were not availabe for use. As it has been shown in figure 7 high maternal blood pressure are associated with both low and normal birth weights.

**Fig 7.**
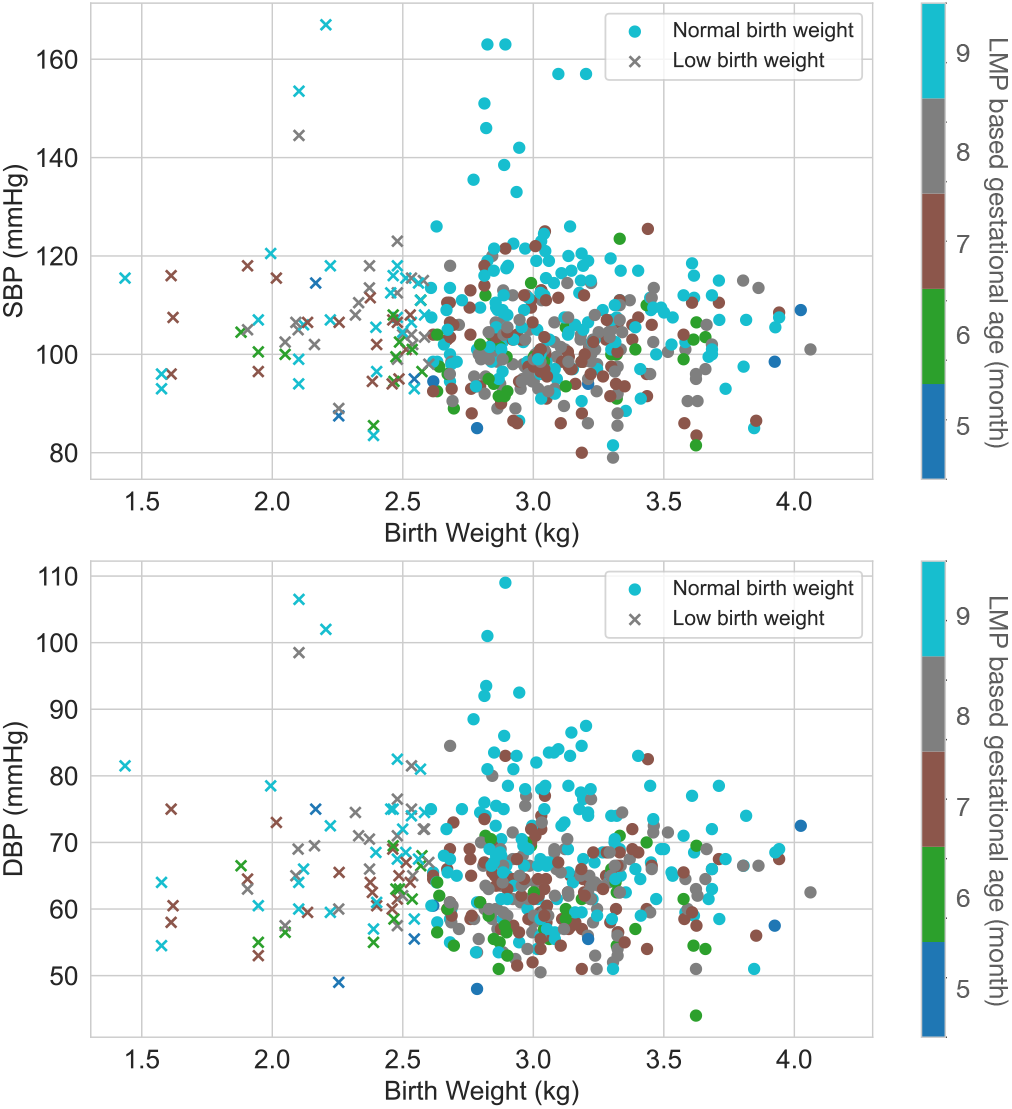
Association of maternal blood pressure and birth weight. Normal Birth Weight is defined by setting weight threshold to 2.64 kg for male newborns, and for females 2.57 kg (30). Different colors indicate gestational age at the time of visit based on last menstrual period (LMP) in month.

Finally, it is important to note that all ultrasound data recorded in this study was not recorded at a specific site, but rather was placed such that a fetal heart beat could be heard. Therefore, the analysis presented here has uncovered changes in the fetal Doppler signal that are related to *maternal* blood flow. This may be due to the relationship between the fetal and maternal cardiovascular systems, or may be due to some other maternal blood flow component interposed between the sensor and the fetus.

## Conclusion

This work presents a novel approach to hypertension detection in pregnancy using a low-cost one-dimensional Doppler probe. To our knowledge, this is the first demonstration of blood pressure estimation from a 1D Doppler device without the need for arterial occlusion / sphygmomanometry. Moreover, since an end-to-end deep learning approach is used, no preprocessing is required to perform the analysis. We demonstrate the utility in a population of 653 pregnant women, with blood pressures in the range of 71 to 169 mmHg SBP and 41 to 110 mmHg DBP, indicating that hypertension is predictable. Interestingly, we noticed differences based on gestational age, which we have also shown can be evaluated from the Doppler device used in this study(31). A dual age-blood pressure prediction therefore can improve performance, and provide a more useful diagnostic.

Notably, we identified this Doppler-hypertension relationship from blood flow on the fetal side of the placenta, indicating that fetal blood flow is significantly affected by high maternal blood pressure. Earlier work on statistical and fluid dynamic models has indicated that there is likely to be a relationship (32–34), and in particular, pathological Doppler velocimetry of the uterine and uteroplacental circulation is a predictor of proteinuric pregnancy-induced hypertension, IUGR and other events in high-risk pregnancies (35–37). Nevertheless, no direct connection between fetal blood pressure and maternal hypertension has been reported in humans before.

The ability to detect hypertensive disorders in pregnancy is particularly important, with few validated devices on the market (26). We note that 1D Doppler recordings are routine in pregnancy, and can be performed at home by the mother or a community healthcare worker (as we demonstrated in (38) using a device that retails at USD$17). By adding this functionality to existing Doppler devices, it presents minimal additional burden and cost to screening and provides the opportunity to identify pre-eclampsia and other hypertension-related disorders during pregnancy and beyond. Only a few seconds of data are required to make an estimate, and estimates can be made on both the fetus and the mother, providing the opportunity to examine blood flow patters around the body in a short period of time.

Moreover, the developed algorithm could run on relatively low cost hardware (such as the Coral TPU), the approach described in this work could be easily implemented in a large range of mHealth and portable devices, leading to rapid and low cost scaling of the technology.

## Data Availability

All data used in this study will be the subject of a future competition and released at that date.

## ACKNOWLEDGEMENTS

The research presented here was funded by a Global Health Grant from Emory University. GC acknowledges the support of the National Institutes of Health, the Fogarty International Center and the Eunice Kennedy Shriver National Institute of Child Health and Human Development, grant number 1R21HD084114-01 (Mobile Health Intervention to Improve Perinatal Continuum of Care in Guatemala), which enabled the collection of the data in this work. GC has financial interest in Alivecor Inc, and receives unrestricted funding from the company. GC also is the CTO of Mindchild Medical ad CSO of Lifebell AI, and has ownership interests in both companies.

## References

1. World Health Organization et al. The World health report: 2005: make every mother and child count. World Health Organization, 2005.

2. Lelia Duley. The global impact of pre-eclampsia and eclampsia. In Seminars in perinatology, volume 33, pages 130–137. Elsevier, 2009.

3. James M Roberts. Preeclampsia: what we know and what we do not know. In Seminars in perinatology, volume 24, pages 24–28. Elsevier, 2000.

4. Michelle Hladunewich, S Ananth Karumanchi, and Richard Lafayette. Pathophysiology of the clinical manifestations of preeclampsia. Clinical Journal of the American Society of Nephrology, 2(3):543–549, 2007.

5. Leona C Poon and Kypros H Nicolaides. Early prediction of preeclampsia. Obstetrics and gynecology international, 2014, 2014.

6. American College of Obstetricians, Gynecologists, et al. Hypertension in pregnancy. report of the american college of obstetricians and gynecologists’ task force on hypertension in pregnancy. Obstetrics and gynecology, 122(5):1122–1131, 2013.

7. Annabelle L Frost, Katie Suriano, Christina YL Aye, Paul Leeson, and Adam J Lewandowski. The immediate and long-term impact of preeclampsia on offspring vascular and cardiac physiology in the preterm infant. Frontiers in Pediatrics, 9:380, 2021.

8. Myung-Kul Yum, Chang-Ryul Kim, Eun-Young Park, and Jong-Hwa Kim. Instability and frequency-domain variability of heart rates in fetuses with or without growth restriction affected by severe preeclampsia. Physiological measurement, 25(5):1105, 2004.

9. Igor Lakhno. Autonomic imbalance captures maternal and fetal circulatory response to pre-eclampsia. Clinical hypertension, 23(1):5, 2017.

10. Igor Lakhno. The impact of preeclampsia on fetal ecg morphology and heart rate variability. Archives of Perinatal Medicine, 20(1):7–10, 2014.

11. Christina YL Aye, Adam J Lewandowski, Pablo Lamata, Ross Upton, Esther Davis, Eric O Ohuma, Yvonne Kenworthy, Henry Boardman, Annabelle L Frost, Satish Adwani, et al. Prenatal and postnatal cardiac development in offspring of hypertensive pregnancies. Journal of the American Heart Association, 9(9):e014586, 2020.

12. Simon Timpka, Corrie Macdonald-Wallis, Alun D Hughes, Nishi Chaturvedi, Paul W Franks, Debbie A Lawlor, and Abigail Fraser. Hypertensive disorders of pregnancy and offspring cardiac structure and function in adolescence. Journal of the American Heart Association, 5(11):e003906, 2016.

13. Glaucimeire Marquez Franco, Marianne de Oliveira Falco, and Waldemar Naves do Amaral. Using ultrasound and doppler ultrasound to assess vascular changes in pre-eclampsia and eclampsia: a systematic review. Reprodução & Climatério, 30(1):33–41, 2015.

14. S Campbell, S Bewley, and T Cohen-Overbeek. Investigation of the uteroplacental circulation by doppler ultrasound. In Seminars in perinatology, volume 11, pages 362–368, 1987.

15. Diana Riknagel, Birthe Dinesen, Henrik Zimmermann, Richard Farlie, Samuel Schmidt, Egon Toft, and Johannes Jan Struijk. Digital auscultation of the uterine artery: a measure of uteroplacental perfusion. Physiological measurement, 37(7):1163, 2016.

16. Dirk Hoyer, Jan Żebrowski, Dirk Cysarz, Hernâni Gonçalves, Adelina Pytlik, Célia Amorim-Costa, Joao Bernardes, Diogo Ayres-de Campos, Otto W Witte, Ekkehard Schleussner, et al. Monitoring fetal maturation—objectives, techniques and indices of autonomic function. Physiological measurement, 38(5):R61, 2017.

17. Yoshua Bengio, Patrice Simard, and Paolo Frasconi. Learning long-term dependencies with gradient descent is difficult. IEEE transactions on neural networks, 5(2):157–166, 1994.

18. Sepp Hochreiter and Jürgen Schmidhuber. Long short-term memory. Neural computation, 9(8):1735–1780, 1997.

19. Dzmitry Bahdanau, Kyunghyun Cho, and Yoshua Bengio. Neural machine translation by jointly learning to align and translate. arXiv preprint 1409.0473, 2014.

20. Kelvin Xu, Jimmy Ba, Ryan Kiros, Kyunghyun Cho, Aaron Courville, Ruslan Salakhudinov, Rich Zemel, and Yoshua Bengio. Show, attend and tell: Neural image caption generation with visual attention. In International conference on machine learning, pages 2048–2057. PMLR, 2015.

21. Prannay Khosla, Piotr Teterwak, Chen Wang, Aaron Sarna, Yonglong Tian, Phillip Isola, Aaron Maschinot, Ce Liu, and Dilip Krishnan. Supervised contrastive learning. arXiv preprint 2004.11362, 2020.

22. Tongzhou Wang and Phillip Isola. Understanding contrastive representation learning through alignment and uniformity on the hypersphere. In International Conference on Machine Learning, pages 9929–9939. PMLR, 2020.

23. Zichao Yang, Diyi Yang, Chris Dyer, Xiaodong He, Alex Smola, and Eduard Hovy. Hierarchical attention networks for document classification. In Proceedings of the 2016 conference of the North American chapter of the association for computational linguistics: human language technologies, pages 1480–1489, 2016.

24. Boris Martinez, Enma Coyote Ixen, Rachel Hall-Clifford, Michel Juarez, Ann C Miller, Aaron Francis, Camilo E Valderrama, Lisa Stroux, Gari D Clifford, and Peter Rohloff. mhealth intervention to improve the continuum of maternal and perinatal care in rural guatemala: a pragmatic, randomized controlled feasibility trial. Reproductive Health, 15(1):120, 2018.

25. Lisa Stroux, Boris Martinez, Enma Coyote Ixen, Nora King, Rachel Hall-Clifford, Peter Rohloff, and Gari D Clifford. An mhealth monitoring system for traditional birth attendant-led antenatal risk assessment in rural guatemala. Journal of medical engineering & technology, 40(7-8):356–371, 2016.

26. Natalie A Bello, Jonathan J Woolley, Kirsten Lawrence Cleary, Louise Falzon, Bruce S Alpert, Suzanne Oparil, Gary Cutter, Ronald Wapner, Paul Muntner, Alan T Tita, et al. Accuracy of blood pressure measurement devices in pregnancy: a systematic review of validation studies. Hypertension, 71(2):326–335, 2018.

27. Samruddhi S Kulkarni, Nasim Katebi, Camilo E Valderrama, Peter Rohloff, and Gari D Clifford. Cnn-based lcd transcription of blood pressure from a mobile phone camera. Frontiers in Artificial Intelligence, 4, 2021.

28. Giancarlo Cicolini, Carmine Pizzi, Elisabetta Palma, Marco Bucci, Francesco Schioppa, Andrea Mezzetti, and Lamberto Manzoli. Differences in blood pressure by body position (supine, fowler’s, and sitting) in hypertensive subjects. American journal of hypertension, 24(10):1073–1079, 2011.

29. C. E. Valderrama, F. Marzbanrad, L. Stroux, B. Martinez, R. Hall-Clifford, C. Liu, N. Katebi, P. Rohloff, and G. D. Clifford. Improving the quality of point of care diagnostics with real-time machine learning in low literacy lmic settings. In ACM SIGCAS Conference on Computing and Sustainable Societies (COMPASS 2018), Jun 2018.

30. Camilo E Valderrama, Faezeh Marzbanrad, Michel Juarez, Rachel Hall-Clifford, Peter Rohloff, and Gari D. Clifford. Estimating birth weight from observed postnatal weights in a guatemalan highland community. Physiological measurement, 41(2):025008, 2020.

31. Camilo E Valderrama, Faezeh Marzbanrad, Rachel Hall-Clifford, Peter Rohloff, and Gari D Clifford. A proxy for detecting iugr based on gestational age estimation in a guatemalan rural population. Frontiers in Artificial Intelligence, 3:56, 2020.

32. A Sengupta, P Biswas, G Jayaraman, and SK Guha. Understanding utero-placental blood flow in normal and hypertensive pregnancy through a mathematical model. Medical and Biological Engineering and Computing, 35(3):223–230, 1997.

33. Aman Gayasen, Sunil Kumar Dua, Amit Sengupta, and D Nagchoudhuri. Effect of non-linearity in predicting doppler waveforms through a novel model. Biomedical engineering online, 2(1):1–13, 2003.

34. Michal Kovo, Jacob Bar, Letizia Schreiber, and Marina Shargorodsky. The relationship between hypertensive disorders in pregnancy and placental maternal and fetal vascular circulation. Journal of the American Society of Hypertension, 11(11):724–729, 2017.

35. P Zimmermann, V Eiriö, J Koskinen, E Kujansuu, and T Ranta. Doppler assessment of the uterine and uteroplacental circulation in the second trimester in pregnancies at high risk for pre-eclampsia and/or intrauterine growth retardation: comparison and correlation between different doppler parameters. Ultrasound in Obstetrics and Gynecology: The Official Journal of the International Society of Ultrasound in Obstetrics and Gynecology, 9(5):330–338, 1997.

36. B Hüneke and Corinna Ude. Uteroplacental and fetal arterial ultrasound doppler flow velocity measurements in unselected pregnancies as a screening test at 32 to 34 gestational weeks. Zeitschrift fur Geburtshilfe und Neonatologie, 206(2):57–64, 2002.

37. Tullia Todros, Ettore Piccoli, Alessandro Rolfo, Simona Cardaropoli, Caterina Guiot, P Gaglioti, Manuela Oberto, Elena Vasario, and I Caniggia. Feto-placental vascularization: a multifaceted approach. Placenta, 32:S165–S169, 2011.

38. B. Martinez, E. Coyote, R. Hall-Clifford, M. Juarez, A. C. Miller, A. Francis, C. E. Valderrama, L. Stroux, G. D Clifford, and P. Rohloff. mhealth intervention to improve the continuum of maternal and perinatal care in rural Guatemala: a pragmatic, randomized controlled feasibility trial. Reproductive Health, In Press, 2018.

